# Urine metabolomic biomarkers linked to C-reactive protein-interleukin-6 axis in persons living with HIV and tuberculosis

**DOI:** 10.1101/2025.08.21.25334178

**Authors:** Andrea B. Doltrario, Myung Hee Lee, Steve Broll, Kathryn Dupnik, Vanessa Rouzier, Patrice Severe, Nancy Dorvil, Jean W Pape, Serena P. Koenig, Daniel W. Fitzgerald, Kyu Y. Rhee

## Abstract

Diagnosing pulmonary tuberculosis (PTB) remains challenging, particularly in people living with HIV (PLWH) who have a high rate of false-negative tests using expectorated sputum. Urine, a non-invasive sample, offers a valuable source of metabolites reflecting systemic changes in disease. This study utilized liquid chromatography–mass spectrometry to investigate urinary biomarkers previously identified in other cohorts, using a well-characterized population of people newly-diagnosed with HIV who screened positive for TB symptoms in Port-au-Prince, Haiti. In this study, we identified a urinary metabolomic signature associated with PTB in PLWH, confirming significant elevations of ureidopropionic acid, 3-hydroxykynurenine, and m/z 115.0498. Untargeted metabolomic analysis revealed a putative isoform of hydroxytryptophan and kynurenic acid as additional PTB-associated metabolites. Four of these five metabolites were also significantly elevated in serum when clinically and microbiologically combined PTB groups were analyzed. Serum metabolite levels correlated positively with elevated blood C-reactive protein (CRP) and IL-6, key inflammatory markers associated with PTB pathology. Moreover, the diagnostic performance of urinary metabolites in participants with CD4+T count below 200 cells/mm³ was not different from that of CRP. Urine metabolomic profiling may complement a patient-centered approach, providing a non-invasive means for TB biomarker discovery and investigating the immunometabolic processes underlying TB in PLWH.

## Introduction

Tuberculosis (TB) diagnosis remains particularly challenging in people living with HIV (PLWH), children, and individuals with extrapulmonary disease (1–3). In these groups, reliance on sputum-based tests (4) is problematic, especially for patients unable to produce sputum or those with paucibacillary pulmonary TB (PTB) (5). More sensitive sputum-based molecular and culture methods exist but are often inaccessible in resource-limited settings (1). Despite World Health Organization (WHO) efforts to promote non-sputum-based diagnostics, existing biomarkers suffer from a high risk of bias and low validation success rates (7), with only a few meeting the WHO Target Product Profile (TPP) accuracy criteria (6,7).

The Abbott Determine™ TB LAM Ag (AlereLAM, USA) is the sole commercial urinary test available, yet its use is restricted by country availability and clinical recommendations, and it demonstrates suboptimal sensitivity (8–10). Despite initial promise, recent evidence shows that first-generation and redesigned LAM assays remain limited in their diagnostic utility (11). Early studies reported FujiLAM sensitivity of 71% in people with HIV (12,13). However, subsequent prospective FujiLAM studies revealed substantial lot-to-lot variability, with important sensitivity and specificity differences (11,14). In a recent evaluation, FujiLAM II demonstrated a sensitivity of only 54% while AlereLAM outperformed it with a sensitivity of 73% (15).

These inconsistencies raise concerns about clinical reliability (11), and while LAM-based diagnostics offer incremental advances, they remain an imperfect alternative for TB diagnosis in PLWH. Urine, a nearly universally accessible biofluid, continues to be a promising point-of-care sample. A recent meta-analysis (10) showed that over 96% of patients could provide urine— regardless of setting—compared with only 69% of inpatients able to produce sputum during the first two days of TB evaluation.

Equally important, urine offers a rich metabolite source that reflects systemic changes during TB infection (16,17). In prior work, we identified and validated ten urinary metabolites elevated in PTB, seven later linked to treatment response and sputum mycobacterial load (18,19). These findings, derived from adult cohorts with few or no PLWH included, led us to hypothesize that some of these metabolites would retain biomarker utility in a symptomatic cohort of antiretroviral therapy (ART)-naïve PLWH in Haiti with a positive TB symptom screen at HIV diagnosis (20). We further hypothesized that urinary metabolites could provide insights into TB immunopathology.

We validated three previously identified urinary metabolites, ureidopropionic acid (UPA), 3-hydroxykynurenine (3-HK), and a compound with m/z 115. 0498, and we identified two additional candidates, with preliminary identification of kynurenic acid (KYNa) and an isoform of hydroxytryptophan. The serum levels of these metabolites correlated positively with key inflammatory markers, including C-reactive protein (CRP), a host-derived molecule frequently proposed as a biomarker for TB (21), and IL-6. Importantly, these urine biomarkers demonstrated diagnostic performance comparable to blood CRP in participants with CD4+ T cell counts below 200 cells/mm³. Urine metabolomic profiling may complement a patient-centered approach, providing a non-invasive means for TB biomarker discovery and investigating the immunometabolic processes underlying TB in PLWH.

## Results

### Previously identified urinary candidate biomarkers of TB were partially validated in PLWH

Our group previously identified elevated levels of ten urinary metabolites (Table 1) in patients with PTB (18). However, the discovery cohort primarily consisted of individuals who did not have HIV, while the control group was asymptomatic. Here, we aimed to validate these findings in a treatment-naïve cohort of PLWH and PTB, using a symptomatic non-TB control group for comparison. The cohort from the study “A Trial of Same-Day Testing and Treatment to Improve Outcomes Among Symptomatic Patients Newly Diagnosed With HIV” (SDART-TB) is described in detail in the Method section. Table 2 provides an overview of the participants’ main characteristics.

**Table 1.** Targeted urinary metabolites and statistical comparison.

| Preliminary identification | Predicted formula | $m/z^A$ | Retention time (min) | $P^B$ |
| --- | --- | --- | --- | --- |
| Unknown 1 | C4H6N2O2 | 115.0498 [M+H]<sup>+</sup> | 1.55 | **0.031** |
| Ureidopropionic Acid (UPA) | C4H8N2O3 | 133.06 [M+H]<sup>+</sup> | 1.56 | **0.028** |
| N1,N12-Diacetylspermine (DiAcSpm) | C14H30N4O2 | 144.1241 [M+2H]<sup>2+</sup> | 14.92 | 0.585 |
| N-Acetylhexosamine | C8H15NO6 | 186.0762 [M+H -2H2O]<sup>+</sup> | 2.08 | 0.411 |
| 3-Hydroxy-kynurenine (3-HK)<sup>C</sup> | C10H12N2O4 | 225.0845 [M+H]<sup>+</sup> | 6.22 | 0.165 |
| Unknown 2 | C9H12N4O4 | 241.0903 [M+H]<sup>+</sup> | 1.77 | 0.225 |
| Neopterin | C9H11N5O4 | 254.0859 [M+H]<sup>+</sup> | 3.32 | 0.524 |
| Sialic Acid 1 | C11H17NO8 | 292.0995 [M+H]<sup>+</sup> | 2.1 | 0.599 |
| Sialic Acid 2 | C11H19NO9 | 310.1148 [M+H]<sup>+</sup> | 2.56 | 0.509 |
| Sialic Acid 3 | C17H26N6O11 | 491.1754 [M+H]<sup>+</sup> | 2.51 | 0.417 |

<sup>A</sup>Mass-to-charge ratio of detected ions. <sup>B</sup>P-values from the Mann-Whitney U test comparing the non-TB group (n = 226) and the microbiologically confirmed TB group (mTB; n = 59). Multiple-comparison adjustment was not applied, as metabolites were pre-specified from prior findings in HIV-negative cohort. A significance threshold of $P < 0.05$ was applied. <sup>C</sup>In an analysis combining the clinically diagnosed TB (cTB) and mTB groups, 3-HK was additionally significantly elevated ( $P = 0.030$ ).

**Table 2.** Characteristics of participants stratified by groups.

|  | **Non-TB (n=226)** | **cTB (n=16)** | **mTB (n=59)** | **p<sup>A</sup>** |
| --- | --- | --- | --- | --- |
| **Age (yr), median [IQR]** | 37.0 [30.3, 45.0] | 40.5 [31.5, 45.0] | 36.0 [31.0, 41.5] | 0.401 |
| **BMI (kg/m<sup>2</sup>), median [IQR]** | 21.0 [19.3, 23.5] | 19.7 [17.1, 21.1] | 18.8 [17.7, 20.5] | <0.001 |
| **Sex, n(%)** |  |  |  | 0.79 |
| Female | 87 (38.5%) | 5 (31.3%) | 24 (40.7%) |  |
| Male | 139 (61.5%) | 11 (68.8%) | 35 (59.3%) |  |
| **HIV status, n(%)** |  |  |  |  |
| Positive | 226 (100%) | 16 (100%) | 59 (100%) |  |
| **CD4+ T count(cells/mm<sup>3</sup>), median [IQR]** | 264 [128, 435] | 197 [62.5, 309] | 167 [86.0, 317] | 0.018 |
| Missing values | 1 (0.4%) | 0 (0%) | 2 (3.4%) |  |
| **CRP (mg/dL), median [IQR]** | 2.56 [0.836, 12.9] | 49.9 [10.2, 126] | 28.0 [4.81, 65.9] | <0.001 |
| **Sputum GeneXpert Ultra<sup>B</sup>, n(%)** |  |  |  | <0.001 |
| Not Detected | 226 (100%) | 16 (100%) | 6 (10.2%) |  |
| Trace |  |  | 6 (10.2%) |  |
| Very Low |  |  | 9 (15.3%) |  |
| Low |  |  | 17 (28.8%) |  |
| Medium |  |  | 15 (25.4%) |  |
| High |  |  | 6 (10.2%) |  |
| **Prevalence of reported symptoms, n(%)** |  |  |  |  |
| Cough | 72 (31.9%) | 14 (87.5%) | 45 (76.3%) | <0.001 |
| Fever | 80 (35.4%) | 10 (62.5%) | 44 (74.6%) | <0.001 |
| Night Sweats | 27 (11.9%) | 5 (31.3%) | 22 (37.3%) | <0.001 |
| Weight Loss | 221 (97.8%) | 15 (93.8%) | 57 (96.6%) | 0.58 |
| **Chest X-ray<sup>C</sup>, n(%)** |  |  |  | <0.001 |
| Abnormal | 13 (5.8%) | 15 (93.8%) | 24 (40.7%) |  |
| Normal | 213 (94.2%) | 1 (6.3%) | 35 (59.3%) |  |

<sup>A</sup> Reported P-values reflect overall comparisons among the three groups using the multigroup statistical test. Numeric variables were tested using ANOVA, and categorical variables were analyzed with the Kruskal-Wallis test for multiple groups. P-values (P) <0.05 were considered statistically significant. <sup>B</sup> Each participant was tested twice with the sputum GeneXpert Ultra. The table reports the highest detected GeneXpert Ultra result per participant. <sup>C</sup> Attending physician chest X-ray assessment. TB: tuberculosis; PTB: pulmonary tuberculosis; cTB: clinical TB group; mTB: microbiological TB group. BMI: body mass index; CRP: C-reactive protein.

Urine samples from 304 participants were initially selected. After excluding three participants whose osmolality values fell below the predefined cutoff of 150 mOsm/kg H₂O, a total of 301 urine samples were analyzed by high-performance liquid chromatography–mass spectrometry (LC-MS), from which data for the 10 metabolites listed in Table 1 were extracted.

The targeted metabolomic analysis revealed that levels of two metabolites– *m/z* 115.0498 (*P* = 0.031) and ureidopropionic acid (UPA; *P =* 0.028) were significantly elevated in PLWH co-diagnosed with PTB when comparing the microbiologically PTB cases (mTB) with the non-TB group, as illustrated in Table 1 and Figure 1A. When using a second comparison category combining participants that were clinically diagnosed with PTB (cTB) and mTB participants, the metabolite 3-hydroxykynurenine (3-HK) was also elevated when compared to the non-TB group (*P* = 0.030), Figure 1B. The described differences remained significant (*P* < 0.05) after adjusting for participants’ characteristics that were different across the groups, such as CD4+T cell count and body mass index (BMI), in a linear regression model (Supplementary Table S1).

**Figure 1.**
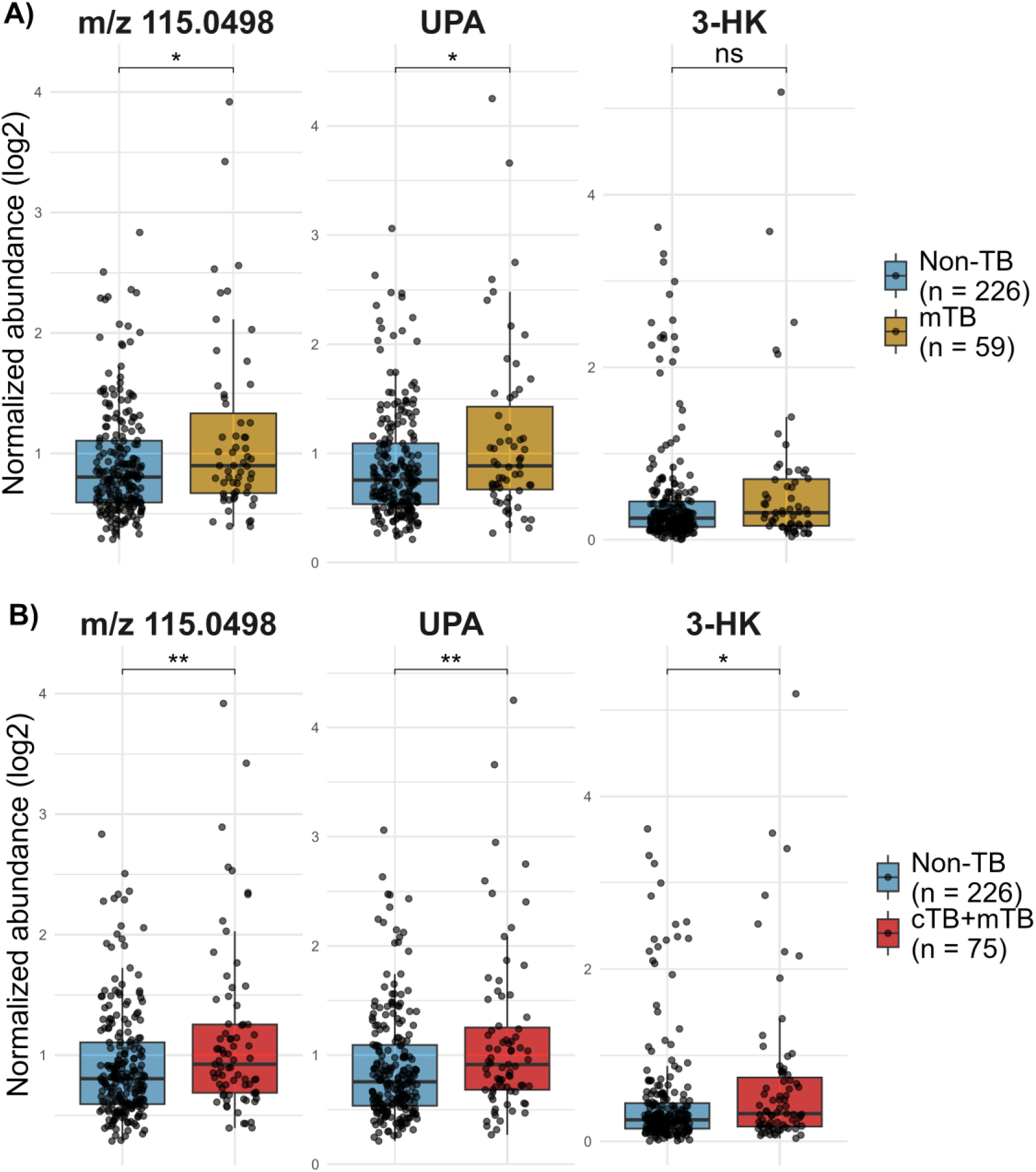
Box Plot comparisons of urinary metabolite abundances between Non-TB and PTB groups. Metabolite abundances are QC median normalized and log₂-transformed. (A) Box plots of m/z 115.0498, UPA, and 3-HK comparing mTB and Non-TB participants. (B) Box plots of m/z 115.0498, UPA, and 3-HK comparing all PTB cases (cTB+mTB) and Non-TB participants. Statistical differences were determined by Mann-Whitney comparison. P-value significance is denoted as follows: *P* < 0.05 (\**), P < 0.01 (**), P < 0.001 (\*\*\**), and ns = not significant. Multiple-comparison adjustment was not applied, as metabolites were pre-specified from prior findings. A significance threshold of P < 0.05 was applied. PTB: pulmonary tuberculosis. UPA: ureidopropionic acid; 3-HK: 3-hydroxykynurenine.

Only the three candidate metabolites described above showed statistically significant differences across the comparisons. The remaining seven metabolites did not exhibit statistically significant differences (Table 1). Regression models from all metabolites from the targeted analysis can be found in Supplementary Table S1.

### Untargeted analysis revealed hydroxy-tryptophan and kynurenic acid as additional candidate biomarkers

Untargeted metabolomic analysis was performed as outlined in the Method section. Six features with the highest posterior probabilities were selected for collision-induced dissociation (CID) and preliminary identification (Table 3). The feature with the highest posterior probability, m/z 222.0956, with a retention time of 5.62 min, was identified as a ^13^C isotope, with the ^12^C isotope having an m/z of 221.0925. With a confidence level of 91% and 84.649% of the total explained intensity, according to Sirius software (22) analysis, this feature was predicted to be a form of hydroxy-tryptophan (HTP). To determine whether this feature corresponded to the known compound 5-HTP, a serotonin precursor derived from tryptophan, we compared it against a 5-HTP pure chemical standard. However, the retention time did not align, indicating that the discovered feature is a different form of HTP (Supplementary Figure S1).

**Table 3.** Top features selected through the untargeted metabolomic pipeline.

| Posterior Probability (%)<sup>A</sup> | Detected ions (m/z) [M+H]<sup>+</sup> | Retention time (min) | Predicted formula | Predicted identification | Details |
| --- | --- | --- | --- | --- | --- |
| 88.3 | 222.0956 | 5.62 | <sup>13</sup>C<sub>11</sub>H<sub>12</sub>N<sub>2</sub>O<sub>3</sub> | Hydroxy-tryptophan (HTP)<sup>B</sup> | HTP <sup>13</sup>C isotope |
| 82.3 | 157.0761 | 5.62 | C<sub>10</sub>H<sub>8</sub>N<sub>2</sub> | Fragment from HTP |  |
| 81.3 | 191.0535 | 1.37 | <sup>13</sup>C<sub>10</sub>H<sub>7</sub>NO<sub>3</sub> | Kynurenic acid (KYNa) | KYNa <sup>13</sup>C isotope |
| 77.6 | 175.0868 | 5.62 | C<sub>10</sub>H<sub>10</sub>N<sub>2</sub>O | Fragment from HTP |  |
| 71.3 | 203.0818 | 5.62 | C<sub>11</sub>H<sub>10</sub>N<sub>2</sub>O<sub>2</sub> | Fragment from HTP |  |
| 66.3 | 190.0503 | 1.37 | C<sub>10</sub>H<sub>7</sub>NO<sub>3</sub> | Kynurenic acid (KYNa) | KYNa <sup>12</sup>C isotope |

The table presents the predicted annotations of the six features with the highest posterior probabilities in the untargeted metabolomics analysis, described by their m/z and retention time values. Four features were identified as potential in-source fragment ions, while two were classified as <sup>13</sup>C isotopes of their predicted parent ions. <sup>A</sup> Posterior probabilities were based on algorithmic confidence and assigned independently of m/z values. <sup>B</sup> Feature m/z 221.0925 was not among the top six and was predicted to be the <sup>12</sup>C isotope of hydroxy-tryptophan.

The additional features with m/z values of 157.0761, 175.0868, and 203.0818, all sharing the same retention time as m/z 222.0956, were putatively assigned as in-source fragments originating from the HTP molecule. These features appear in lower abundances (Supplementary Figure S1) than the parent ion, and they are predicted to exist as fragments of hydroxytryptophan isoforms when analyzed using Competitive Fragmentation Modeling for Metabolite Identification tool (23) based on Simplified Molecular Input Line Entry System (SMILES) format queries.

The feature with m/z 191.0535 and retention time of 1.37 minutes was identified as the ^13^C isotope from the ^12^C feature with m/z of 190.0503. By matching its retention time and fragmentation pattern with those of a pure chemical standard, the feature was predicted to be kynurenic acid (KYNa), as illustrated in Figure 2.

**Figure 2.**
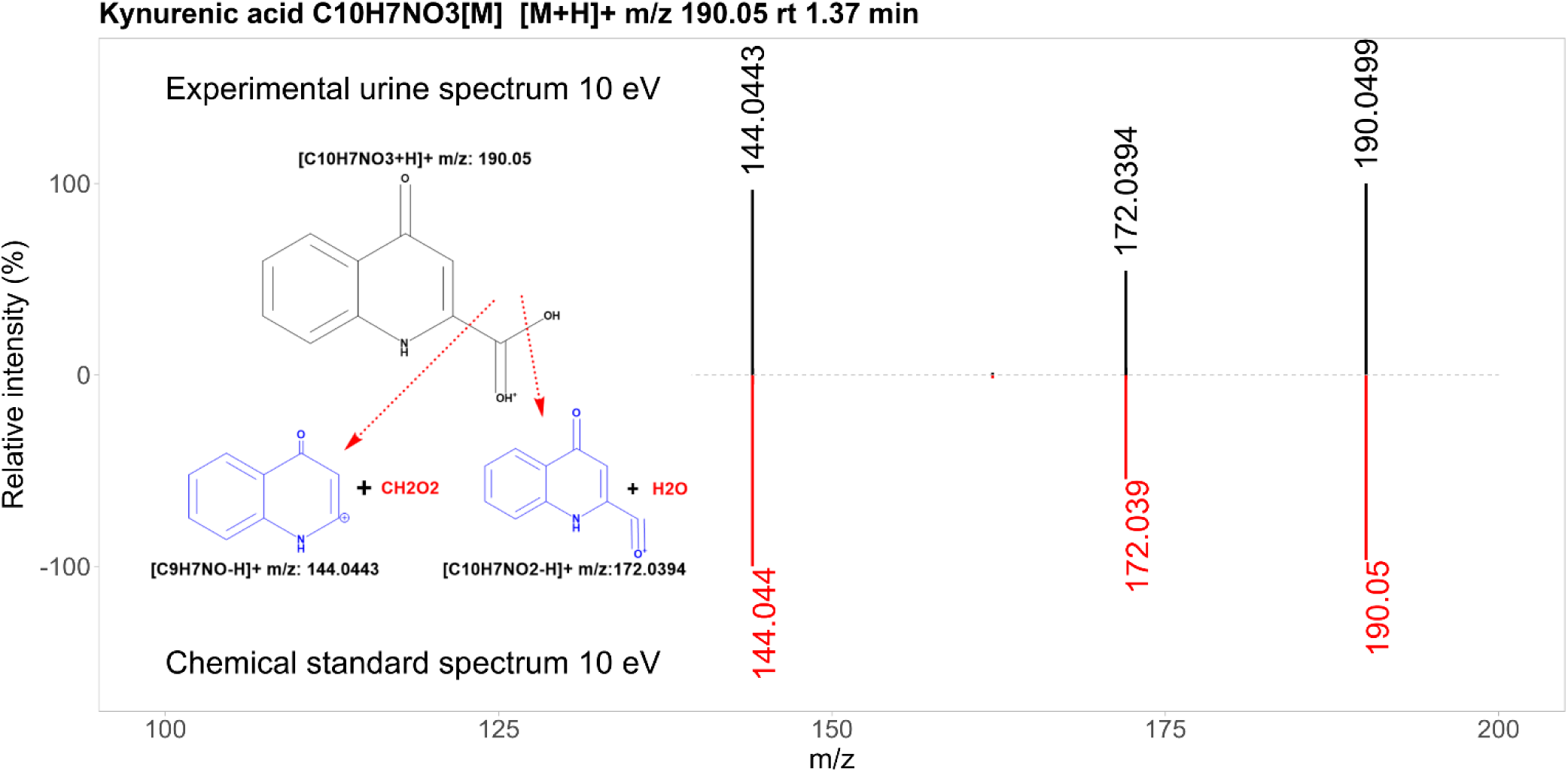
Mirror plot of experimental urine and chemical standard fragmentation spectra for kynurenic acid (KYNa). The plot depicts the most relevant fragments obtained from the collision-induced dissociation at 10 eV. Fragmentation was performed on urine samples and a pure chemical standard. The mass-to-charge ratio (m/z) and relative intensity of the fragments match between urine and pure chemical standard, supporting the identification of the metabolite as kynurenic acid.

### Serum metabolomic analyses reveal complementary information to urine

To complement the investigation of urinary candidate biomarkers and their association with PTB-related inflammatory markers such as CRP and IL-6, we conducted a pilot analysis using serum samples within the SDART-TB cohort. Metabolomic profiling and cytokine quantification were performed on these samples. The design of this cohort subset is described in detail in the Methods section. Briefly, the pilot included 80 participants: 16 with clinically diagnosed TB (cTB), 32 with microbiologically confirmed TB (mTB), and 32 non-TB controls. Serum metabolite abundances were extracted for the three targeted metabolites, which were differentially abundant in urine, and the two additional features identified through untargeted analysis. Supplementary Table S2 provides an overview of the subset participants’ main characteristics.

We first noticed that, although differences were not statistically significant, cTB patients consistently showed the highest median levels of m/z 115.0498, UPA, and 3-HK compared to the mTB group. In serum, within the subset of participants, cTB patients showed higher median levels of m/z 115.0498 (0.844 vs. 0.763 in mTB, *P=* 0.269), UPA (0.727 vs. 0.476, *P=* 0.216), and 3-HK (1.071 vs. 0.972, *P=* 0.477). This trend was also observed for the urinary levels, where, cTB participants exhibited elevated normalized abundances of m/z 115.0498 (1.091 vs. 0.865 in mTB, *P=* 0.433), UPA (1.009 vs. 0.848, *P=* 0.441), and 3-HK (0.418 vs. 0.243, *P=* 0.184). The non-significant p-values may be due to the small sample size of the cTB group. A larger cohort of clinically diagnosed PTB participants would be needed to validate this finding. This trend was not observed for metabolites identified through untargeted analysis, likely due to methodological differences in detection and quantification compared with targeted metabolites.

Second, we compared the PTB participants with the non-TB controls. When comparing the mTB group to the non-TB group, most metabolites in serum visually exhibited an upward trend, as shown in Figure 3A, but only HTP was significantly elevated (*P* = 0.0052). The features m/z 115.0498, UPA, KYNa, and 3-HK did not show statistically significant differences, with *P*-values of 0.1281, 0.0948, 0.9947, and 0.0948, respectively.

**Figure 3.**
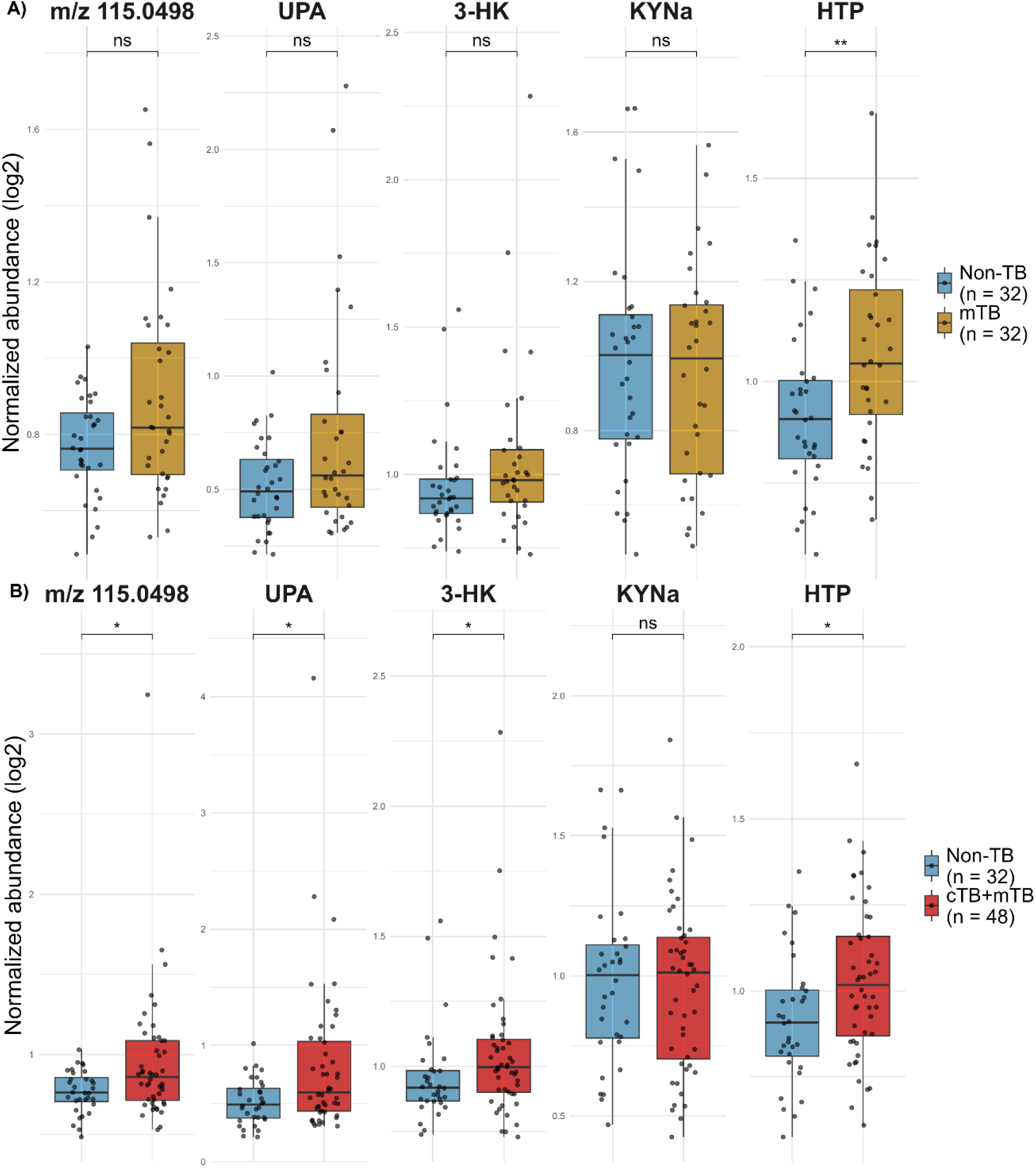
Box Plot comparisons of serum metabolite abundances between PTB and Non-TB groups. Metabolite abundances are QC median normalized and log₂-transformed. (A) Box plots of m/z 115.0498, UPA, 3-HK, KYNa, and HTP comparing mTB and Non-TB participants. (B) Box plots of m/z 115.0498, UPA, 3-HK, KYNa, and HTP comparing all TB cases (cTB+mTB) and Non-TB participants. Statistical differences were determined by Mann-Whitney comparison. P-value significance is denoted as follows: *P* < 0.05 (\**), P < 0.01 (**), P < 0.001 (\*\*\**), and ns = not significant. Multiple-comparison adjustment was not applied. A significance threshold of P < 0.05 was applied. PTB: Pulmonary tuberculosis; UPA: ureidopropionic acid; 3-HK: 3-hydroxykynurenine. KYNa: kynurenic acid; HTP: Hydroxy-tryptophan.

When combining cTB and mTB groups (Figure 3B), four out of five metabolites exhibited significant elevation in the PTB group compared to the control group: m/z 115.0489 (*P* = 0.0338), UPA (*P* = 0.0157), 3-HK (*P* = 0.0373), and HTP (*P* = 0.0186). KYNa, however, once again showed no significant difference between the groups (*P* = 0.9106).

It is worth noting that serum levels of 3-HK were generally low, with abundances approaching the lower detection limit for the analytical method. Moreover, after correcting for BMI, 3-HK abundances were not significantly elevated in the PTB group using the second reference category (cTB and mTB combined). This finding aligns with existing literature indicating that kynurenine 3-monooxygenase (KMO)—the enzyme responsible for catalyzing the production of 3-HK—is highly expressed in peripheral tissues such as the kidney (24). This tissue-specific expression may explain and the statistical findings and the higher levels of 3-HK detected in urine compared to serum.

### Both blood C-reactive protein (CRP) and IL-6 levels are elevated in PTB patients

Blood CRP levels in participants from the SDART-TB clinical trial have been previously reported (21). The present study extends these findings by reporting CRP levels in a subset of selected control and PTB participants who met our inclusion criteria, showing elevated CRP in both PTB categories (Figure 4A and 4B). Given that IL-6 is the primary stimulator of CRP production, we also examined whether IL-6 levels were elevated in serum.

**Figure 4.**
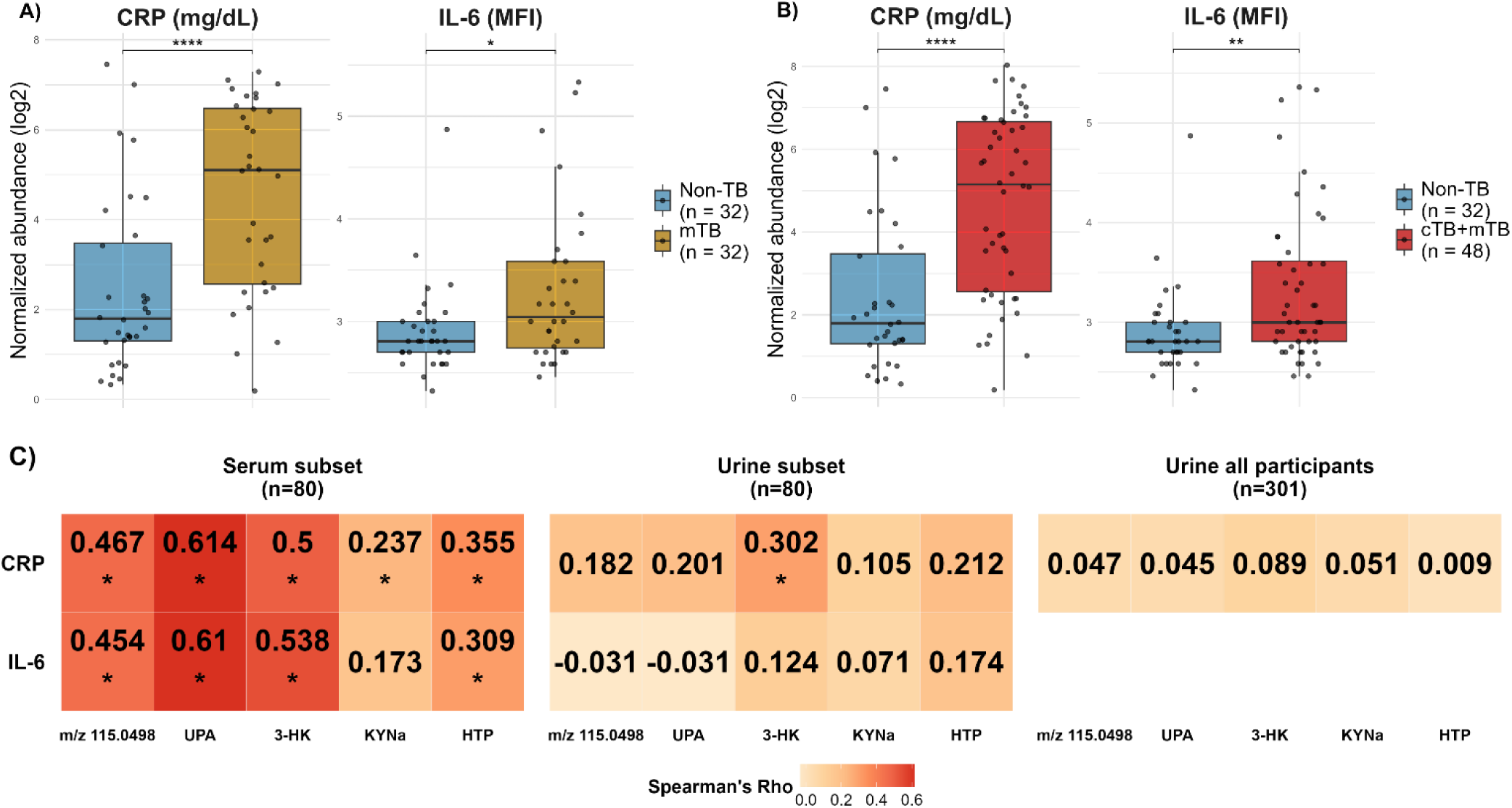
Box Plot comparisons of CRP and IL-6 abundances between Non-TB groups and PTB and their correlation with metabolites. (A) Box plots of blood CRP and IL-6 serum abundances comparing mTB and Non-TB participants. (B) Box plots of blood CRP and IL-6 serum abundances comparing all TB cases (cTB+mTB) and Non-TB participants. Statistical differences in (A) and (B) were determined by Mann-Whitney comparison. P-value significance is denoted as follows: *P* < 0.05 (\**), P < 0.01 (**), P < 0.001 (\*\*\**), and ns = not significant, representing unadjusted values from the statistical tests. (C) Heatmap of Spearman correlation coefficients between urinary metabolites and inflammatory markers across different TB subsets. Each value in the heatmap represents the Spearman correlation coefficient (ρ). An asterisk (*) indicates statistically significant correlations after Holm–Bonferroni correction for multiple testing (*P* < 0.05). PTB: Pulmonary tuberculosis; CRP: C-reactive protein; MFI: Mean fluorescence intensity; UPA: ureidopropionic acid; 3-HK: 3-hydroxykynurenine; KYNa: kynurenic acid; HTP: Hydroxy-tryptophan.

As shown in Figure 4A and 4B, median IL-6 levels were significantly higher in the PTB group compared to non-TB controls for both PTB categories: mTB alone (*P* = 0.0219) and cTB and mTB combined (*P* = 0.006). Furthermore, within the Th1/Th2 cytokine panel tested, no other cytokine exhibited significant differences between the groups (Supplementary Table S3). We also identified that the quantified serum IL-6 levels positively correlated with the blood CRP levels (rho=0.70, *P*<0.001).

### Serum metabolite levels correlate positively with CRP and IL-6 levels

Next, we aimed to determine if the abundances of key metabolites in serum and urine correlated with CRP and IL-6 levels. The correlation analysis was performed on both the subset and the full cohort, without stratifying participants into control or PTB subgroups. By analyzing all participants together, we aimed to preserve statistical power and better capture global associations with CRP and IL-6 across the full spectrum of values.

Figure 4C presents a heatmap of the correlation analysis and shows that all serum metabolites exhibited statistically significant positive correlations with CRP levels. Similarly, all serum metabolites, except for KYNa (rho = 0.24, Adj. *P* = 0.124), also showed significantly positive correlations with IL-6 levels (Figure 4C). Notably, when performing the correlation for urinary metabolites for this same subset of participants, no significant correlation was observed between urine metabolite levels and IL-6. Only urine levels of 3-HK demonstrated a positive correlation with CRP levels (rho = 0.30, Adj. *P* = 0.0337). In a broader analysis involving the whole cohort, with 301 participants, no significant correlations were observed between urine metabolites and CRP levels (Figure 4C). The correlation with IL-6 was not performed for the full cohort since IL-6 levels were only available for a subset of participants.

Distinct correlation patterns emerged between serum and urinary metabolites with CRP and IL-6. Consistent with our findings, Schlosser et al. (2023) (25) reported that plasma and urine provide complementary metabolic information, with certain genetic associations unique to each biofluid. These results indicate that paired analyses can reveal correlations with clinical markers that may be overlooked in a single matrix, allowing us to capture systemic and kidney-specific processes.

### Urinary metabolites had modest overall diagnostic performances but were comparable to CRP in severe immunosuppression

We evaluated the diagnostic performance of urinary candidate biomarkers in this cohort. The area under the curve (AUC) of Receiver Operating Characteristic (ROC) curves was determined for the three targeted metabolites, the two untargeted metabolites, and CRP. These calculations were performed for both reference categories, mTB alone and cTB and mTB combined. As shown in Table 4, in the overall analysis (with all participants), urinary metabolites demonstrated modest diagnostic performance, with m/z 115.0498, UPA, and 3-HK achieving the highest AUC values (60.3%, 60.7%, and 58.3% respectively) when combining cTB and mTB groups. CRP exhibited a superior performance of 75.8% for the same comparison.

**Table 4.** Comparative diagnostic performance of urinary metabolites and blood CRP.

|  | All participants | CD4+ T cell count<sup>A</sup> |  |
| --- | --- | --- | --- |
|  |  | <200 cells/mm<sup>3</sup> | >200 cells/mm<sup>3</sup> |
|  |  | ROC AUC (%) [ 95% CI ] |  |
| **mTB vs non-TB** | **n=285** | **n=117** | **n=165** |
| m/z 115.0498 | 59.1 [50.8, 67.3] * | 62.9 [51.6, 74.1] | 53.5 [40.3, 66.6] * |
| UPA | 59.3 [50.9, 67.6] * | 63.3 [52.2, 74.5] | 54.4 [41.2, 67.7] * |
| 3-HK | 55.9 [47.2, 64.6] * | 64.9 [53.8, 76.0] | 45.4 [31.7, 59.2] * |
| KYNa | 60.3 [52.8, 67.8] * | 65.5 [54.8, 76.2] | 56.3 [45.3, 67.4] * |
| HTP | 55.5 [47.5, 63.5] * | 65.2 [53.6, 76.7] | 45.4 [34.8, 56.0] * |
| Blood CRP | 74.6 [67.6, 81.6] | 70.8 [60.2, 81.4] | 74.1 [62.9, 85.3] |
| **cTB + mTB vs non-TB** | **n=301** | **n=125** | **n=173** |
| m/z 115.0498 | 60.3 [53.1, 67.6] * | 64.2 [54.1, 74.3] | 55.4 [44.2, 66.6] * |
| UPA | 60.7 [53.4, 68.1] * | 64.8 [54.8, 74.9] | 56.2 [45.0, 67.4] * |
| 3-HK | 58.3 [50.6, 66.0] * | 67.4 [57.3, 77.4] | 51.5 [39.6, 63.3] * |
| KYNa | 59.2 [52.1, 66.3] * | 64.8 [54.6, 75.0] | 54.6 [44.3, 64.8] * |
| HTP | 55.0 [47.6, 62.3] * | 64.4 [53.8, 74.9] | 54.3 [44.3, 64.3] * |
| Blood CRP | 75.8 [69.6, 82.0] | 71.1 [61.5, 80.8] | 76.7 [67.2, 86.1] |

Area under the ROC curve (AUC) values and their confidence interval (CI) are shown for urinary metabolites and blood CRP in distinguishing PTB from non-TB controls. Comparisons between urinary metabolites and CRP performances within each group were assessed using DeLong's test, with $P < 0.05$ considered statistically significant. An asterisk (\*) indicates a significant DeLong test result. Notably, the performance of urinary metabolites did not differ from that of CRP among participants with CD4<sup>+</sup> T cell counts < 200 cells/mm<sup>3</sup>. <sup>A</sup>Three participants who did not have CD4<sup>+</sup> T cell count values were excluded from the CD4<sup>+</sup> T cell count subgroup stratification. ROC: receiver operating characteristic; CRP: C-reactive protein; UPA: ureidopropionic acid; 3-HK: 3-hydroxykynurenine; KYNa: kynurenic acid; HTP: Hydroxy-tryptophan; mTB: microbiological TB group; cTB: clinical TB group.

KYNa and HTP achieved their highest diagnostic performance when using the microbiological reference category (mTB) with an AUC of 60.3% and 55.5%, respectively.

DeLong’s test (26) was used to compare AUCs between biomarkers within each group. Non-overlapping confidence intervals and DeLong’s test results (where *P* < 0.05 indicates significantly different AUCs) showed that CRP outperformed urinary metabolites in the overall cohort for both reference categories—mTB alone and the combined cTB and mTB groups (Table 4).

We further investigated whether diagnostic performance varied when stratifying participants by CD4+ T-cell count, using the well-established threshold of 200 cells/mm³ to define severe immunosuppression in PLWH. In the subset with CD4⁺ T cell counts > 200 cells/mm³, like for the overall cohort, we observed that urinary metabolites performed significantly worse than CRP. However, and interestingly, urinary metabolites performed as well as CRP in participants with CD4 < 200 cells/mm³, combining cTB and mTB participants, or using mTB alone. In this same subset of participants, urinary metabolites demonstrated better AUC performance than in the overall cohort, unlike blood CRP, which showed no improvement in performance.

## Discussion

In this study, we identified a urinary metabolomic signature associated with PTB in PLWH, confirming significant elevations of ureidopropionic acid, 3-hydroxykynurenine, and m/z 115.0498. Untargeted analysis additionally revealed an isoform of hydroxytryptophan and kynurenic acid as PTB-associated metabolites. Four of these five metabolites were also significantly elevated in serum when analyzing cTB and mTB groups combined and correlated positively with key inflammatory markers, CRP, and IL-6. Moreover, the diagnostic performance of urinary metabolites in participants with advanced immunosuppression was not different from that of CRP.

Two significantly elevated metabolites, 3-HK and KYNa, are produced downstream from tryptophan degradation via the kynurenine pathway (Figure 5). This elevation is consistent with previous studies showing increased kynurenine metabolites, including kynurenine and 3-HK, during Mtb infection in numerous metabolomic studies, using both human biospecimens and in vitro models (27–34). Stone et al. (2024) (35) further noted that KYNa can arise not only through classical kynurenine aminotransferase (KAT) activity but also via an alternative IL4-induced protein (IL4i) pathway producing indole pyruvate, which cyclizes into KYNa.

**Figure 5.**
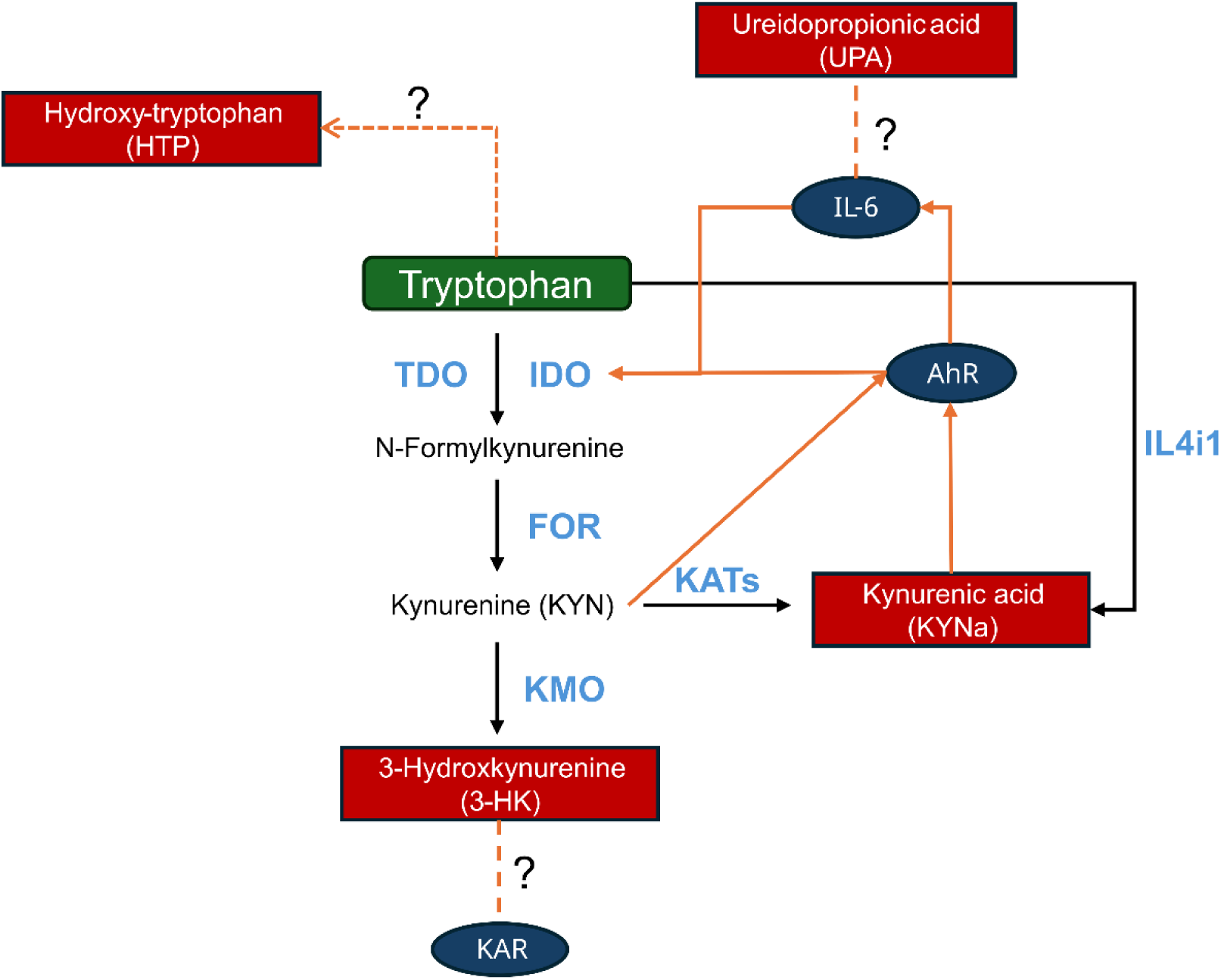
Proposed schematic of tryptophan degradation through the kynurenine pathway and its potential interactions in TB immunopathology. This figure illustrates a schematic of the kynurenine pathway and its possible links to other immune markers and TB immunopathology. Tryptophan is highlighted in a green box. Candidate biomarkers found elevated in PTB urine samples are highlighted in red boxes. Enzymes involved in the pathway are shown in light blue. Black arrows indicate established biochemical reactions, while orange arrows depict proposed relevant additional interactions to TB immunopathology. Dashed orange lines represent hypothetical connections between components that require further investigation. IDO: Indoleamine 2,3-dioxygenase; TDO: Tryptophan 2,3-dioxygenase; KAT: Kynurenine aminotransferase; FOR: Formamidase; KMO: Kynurenine 3-monooxygenase; KAR: Kainate receptor; AhR: Aryl hydrocarbon receptor; IL4i1: Interleukin 4 Induced 1.

Although Indoleamine 2,3-dioxygenase (IDO) activation reduces host tryptophan availability, Mtb can synthesize its own, suggesting that host-driven depletion may not impair bacterial survival or host outcome (36). Instead, accumulated kynurenine metabolites may support immune evasion by suppressing T cell activity and facilitating persistent infection (29,37), which could be exploited by *Mycobacterium tuberculosis* (Mtb). 3-HK may additionally have concentration-dependent, pleiotropic roles in infection. Parada-Kusz et al. (2024)(38) recently identified 3-HK as a kainic acid receptor (KAR) antagonist, enhancing host survival in zebrafish models of lethal Gram-negative infections. While its direct protective effects in Mtb remain untested, the consistent finding of its upregulation in PTB suggests a role in immune modulation and disease severity.

Also derived from tryptophan (Figure 5), our untargeted analysis identified a hydroxytryptophan (HTP) isomer as a complementary PTB marker for this cohort. The detected HTP isomer did not match 5-HTP, suggesting a distinct structure of potential biological relevance. HTP isomers with a different retention time from 5-HTP are reported to be found in urine (39). Additionally, tryptophan hydroxylation can occur at multiple carbon positions through enzymatic and non-enzymatic mechanisms (40,41). We hypothesize that this HTP isomer may arise either enzymatically or from non-enzymatic hydroxylation driven by inflammation and reactive oxygen species (ROS) (42). The Fenton reaction can chemically hydroxylate tryptophan at the 4-, 5-, 6-, and 7-positions (43), and in TB, elevated labile iron enhances ROS production via this reaction, generating hydroxyl radicals that contribute to bacterial control and tissue pathology (44,45).

In this cohort, PTB patients showed elevated CRP and its primary inducer, IL-6. Serum metabolites correlated positively with these established inflammatory markers. CRP is a well-studied TB biomarker, and it is typically elevated in TB regardless of HIV status (46,47). IL-6 plays dual roles, mediating immune responses against Mtb while also contributing to immunosuppression under certain conditions (48–50). Its elevation has been consistently observed in TB patients and in vitro infection models (49,51–54).

Losada et al. (2020) (52) reported higher IL-6 levels in bronchoalveolar lavage fluid (BALF) from smear-negative PTB cases with greater radiological severity. In this cohort, we found that the median level of CRP in blood from patients in the cTB group was higher than that of the mTB group. The cTB group also had the highest prevalence of chest X-ray abnormalities (93.8% in cTB vs. 40.7% in mTB) and reported cough (87.5% in cTB vs. 76.3% in mTB) and higher levels of m/z 115.0498, UPA, and 3-HK in urine and serum. Based on these findings, patients in the cTB group appeared to exhibit more severe clinical presentations overall. In contrast, clinical severity likely varied among individuals in the mTB group, as diagnoses had a microbiological confirmation, regardless of symptom severity. We therefore hypothesize that differences in disease severity may explain the metabolite variations observed between the cTB and mTB groups.

We further hypothesize that these metabolites are linked to IL-6’s role in TB pathogenesis. IL-6 activates IDO (35), converting tryptophan into kynurenine, an endogenous AhR ligand (55). AhR activation, in turn, enhances IL-6 transcription, perpetuating this inflammatory cycle (56) (Guarnieri, 2020). Liu et al. (2024) (57) showed that the kynurenine–aryl hydrocarbon receptor (AhR) loop delays T-cell responses in the lungs during TB infection.

KYNa, elevated in PTB cases in this study, may contribute to the AhR–immune regulation loop, as it is an even more potent AhR agonist (35,58). Supporting AhR’s role in immune responses during Mtb infection, Memari et al. (2015) (59) showed strong AhR activation in Mtb-infected macrophages, enhancing immune-regulatory gene expression. AhR also detects bacterial virulence factors such as Mtb’s phthiocol (60) and interacts with TB drugs like rifabutin and rifampicin (61).

Ureidopropionic acid has also been linked to IL-6 regulation. Luo et al. (2024) (62) reported that UPA reduced IL-6 and IL-1β mRNA in LPS-stimulated RAW264.7 cells, suggesting an anti-inflammatory role. In our study, we found a positive correlation between UPA and IL-6, potentially reflecting a negative feedback mechanism or differences from the macrophage model. UPA’s clinical relevance in TB was independently highlighted in TB meningitis by another group that demonstrated its cerebrospinal fluid levels correlated with higher mortality (63).

We also compared candidate biomarkers across biofluids for a subset of participants. Serum m/z 115.0498, UPA, 3-HK, and HTP followed the same trend as in urine, with elevated levels in PTB cases. Statistically, HTP showed a statistically significant result when comparing only the mTB group with controls. When combining cTB and mTB groups, the elevation of metabolites was statistically significant, except for KYNa. The authors hypothesize that cTB contributes to higher levels of metabolites and increases sample size. In contrast, the first reference category of comparison (mTB alone), likely due to lower levels and the smaller sample size, had reduced statistical power.

Interestingly, unlike serum, urine metabolite abundances did not correlate with CRP in the overall cohort, and only 3-HK showed a potential positive correlation in the subset analysis. This suggests that additional factors influence urinary metabolite levels beyond their systemic concentrations. Urine may also capture localized metabolite production in affected tissues, sufficient to increase excretion without markedly altering serum levels. Notably, urine analysis revealed significant KYNa differences between control and PTB groups that were absent in serum, indicating that urine may be a superior biofluid for biomarker detection, reflecting both systemic and tissue-specific pathology.

Not all previously identified metabolites were elevated in this cohort, likely reflecting differences in population characteristics. While both this study and the 2018 Isa et al.(18) study included participants from Haiti, they differed markedly in HIV status and clinical features. Isa et al.’s cohort was predominantly HIV-negative (only 15% were PLWH), whereas the present study exclusively enrolled PLWH. None of the participants in either study were receiving ART at enrollment. Moreover, Isa et al. included asymptomatic controls, whereas all participants here—controls included—were symptomatic. CD4+T cell counts were also higher in Isa et al.’s cohort (433–536 cells/mm³) compared to the SDART cohort (167–264 cells/mm³), indicating more advanced immunosuppression in this study. Sex distribution was comparable.

HIV independently upregulates IDO activity, driving persistent inflammation even without Mtb infection (27,64). CD4+ T cell depletion weakens immune defenses and disrupts gut integrity, promoting microbial translocation and chronic immune activation, eventually leading to immune exhaustion (64). These overlapping inflammatory processes may explain the partial validation of candidate biomarkers in this cohort, as metabolite changes could reflect a broader inflammatory state in PLWH. Other infections, such as bacterial pneumonia or opportunistic infections, may also contribute to altered metabolite profiles.

The authors acknowledge limitations. Particularly in PLWH and despite extensive screening for extrapulmonary TB, undetected sites of infection may have influenced metabolite profiles. Biomarker development is further complicated by the dynamic nature of metabolites during disease progression. Participants were enrolled at the time of HIV infection diagnosis, but this does not differentiate between recent and longstanding HIV or Mtb infections.

As with many biomarker studies, this work relied on biobanked samples originally collected for other purposes, which can constrain sample size and clinical data availability. Additionally, the urine metabolome is influenced by factors such as age, sex, BMI, diet, microbiota, genetics, and hydration status (65–67), complicating standardization. Technical limitations, including chromatographic resolution and data processing, may also obscure subtle changes in metabolite levels (68–71).

Despite these challenges, this study provides key biological insights, revealing urinary UPA, 3-HK, KYNa, and a hydroxytryptophan isomer as a metabolomic signature in PLWH with PTB that links host immune regulation and metabolic reprogramming. UPA may be linked to IL-6 regulation, kynurenine pathway metabolites (3-HK, KYNa) reflect IDO activation and possible AhR-driven immune modulation, and the hydroxytryptophan isomer suggests oxidative stress or enzymatic tryptophan hydroxylation. Importantly, urinary metabolites performed comparably to blood CRP in participants with CD4 counts <200 cells/mm³. Thus, demonstrating diagnostic potential in patients with severe immunosuppression. These findings pave the way for deeper exploration of metabolite-driven interactions in TB pathogenesis and host immune modulation, emphasizing their role as active mediators of disease establishment and progression.

## Methods

### Clinical Cohort

The researchers worked with urine samples collected from participants enrolled in the study “A Trial of Same-Day Testing and Treatment to Improve Outcomes Among Symptomatic Patients Newly Diagnosed With HIV” (SDART-TB) (clinicaltrials.gov ID NCT03154320). Results from the trial are detailed previously (20). Briefly, individuals newly diagnosed with HIV, aged ≥18 years, not currently receiving TB treatment, or pregnant, were screened for participation. Enrollment was conducted between 2017 and 2020 at the GHESKIO Center (Haitian Group for the Study of Kaposi’s Sarcoma and Opportunistic Infections) in Port-au-Prince, Haiti.

Participants reporting at least one symptom of TB—cough, fever, night sweats, or weight loss—were invited to join the study, and 500 met inclusion criteria (excluding those who declined participation, were not ready to accept their HIV diagnosis, or had severe comorbidities).

At enrollment, clinical and demographic data were collected, including age, sex, BMI, household income, clinical symptoms, and baseline CD4⁺ T-cell count. Additional laboratory measures, such as CRP levels, sputum Xpert MTB/RIF Ultra results, and chest X-ray findings, were also obtained. Bacteriological screening for PTB using Xpert Ultra and sputum culture was conducted at baseline, and diagnoses of bacteriologically confirmed or empirically treated TB were recorded accordingly. Importantly, all samples were collected prior to the initiation of anti-TB therapy and ART.

The microbiological TB (mTB) group included participants with at least one positive sputum-based bacteriological test (Xpert Ultra or sputum culture). The clinical TB (cTB) group consisted of participants who screened positive for symptoms, had negative sputum bacteriological results, but were initiated on empirical treatment for PTB based on high clinical suspicion. The non-TB control group consisted of participants who screened positive for symptoms but had negative sputum bacteriological results and were not diagnosed with pulmonary TB following medical evaluation.

The SDART-TB cohort has also been studied in relation to CRP (21), PTB predictors (72), and co-infection with Histoplasma (73).

### Sex as a biological variable

The SDART-TB clinical trial and the current metabolomic study included participants of both sexes. Efforts were made to ensure proportional representation of male and female participants in both the PTB and control groups.

### Study Design

The present study was conducted within the SDART-TB clinical cohort to evaluate urinary metabolomic biomarkers associated with PTB, with an additional pilot study focusing on serum cytokine and metabolite profiling.

From the initial pool of 500 participants enrolled in the SDART-TB cohort, we first identified those with urine samples collected at enrollment (n = 460) for inclusion in the present metabolomic study. From those, all participants with PTB, defined as either cTB (n = 16) or mTB (n = 60), were included. To achieve a 3:1 control-to-case ratio, control participants were selected to maintain comparable frequencies of sex and age across the PTB (n = 76) and control (n = 228) groups. Table 2 summarizes the main demographic and clinical characteristics of the 304 selected participants.

A pilot study was designed to evaluate both cytokine and metabolite levels in serum. Participants with sufficient serum volume for both assays were first identified, and all available clinically diagnosed TB cases meeting this criterion were included. To maximize representation of the cTB group, which exhibited the highest blood CRP levels in the parent cohort, we selected participants while balancing for age and sex across groups. The final serum pilot subset included 16 cTB, 32 mTB, and 32 non-TB participants.

### Sample Preparation for LC-MS

Urine samples were collected in Haiti and frozen at −80°C and shipped on dry ice to New York for long-term storage at −80°C. Urine sample preparation and data acquisition were as previously described (18,19). Briefly, 300 μL of urine was centrifuged for 2 min, at 4°C at 15,000 rpm to remove pelleted debris, followed by filtration with a PALL nanosep device (3k omega filter) to eliminate remaining large particles. Urine samples were normalized to 150 mOsm/kg H₂O prior to data acquisition. Osmolality was measured using an Advanced Instruments Model 2020 osmometer, and dilutions were performed with Milli-Q water. Following filtration and dilution, samples were mixed in a 1:1 ratio with acetonitrile containing 0.2% formic acid and centrifuged at 15,000 rpm for 8 minutes at 4 °C. Finally, 100 µL of the clarified supernatant was loaded into LC-MS vials for metabolomic analysis.

Serum samples were collected in Haiti and frozen at −80°C before being shipped on dry ice to NY for long-term storage at −80°C. Aliquots were thawed on ice and mixed thoroughly before proceeding with sample preparation. To remove protein, a 100 µL serum aliquot was combined with methanol (2:1 serum: methanol ratio) and incubated at 4°C with shaking at 300 rpm for 10 minutes. The mixture was centrifuged at 15,000 rpm for 10 minutes at 4 °C. Supernatant was mixed with acetonitrile+0.2% formic acid in a 1:1 ratio and centrifuged at 15,000 rpm for 10 minutes at 4 °C. Finally, 100 µL of the clarified supernatant was collected and loaded into LC-MS vials for metabolomic analysis.

### Data Acquisition and Processing

Urine samples were processed in duplicates for the LC-MS experiments in sets containing 25 or 26 blinded and randomized samples. Quality control (QC) samples were prepared by pooling an equal volume of urine samples from 10% of the total cohort, ensuring representation from all study groups. The same pooled QC sample was included in each analytical batch. Data acquisition was performed using an Agilent Accurate Mass 6546 QTOF (quadrupole time-of-flight) spectrometer with Formic Acid method as described elsewhere (18,19). Detected ions were indexed and characterized using their ion m/z and chromatographic retention time. Data were analyzed using Agilent Technologies Qualitative Analysis B.10 (74), Agilent Technologies MassHunter Profinder B.10 (75), and XCMS software (76). Targeted metabolites were verified by running their respective chemical standards with each set of urine samples. The following pure chemicals were used for the chemical standards: ureidopropionic acid (product #94295, Sigma Aldrich), neopterin (product #N3386, Sigma Aldrich), n-acetylneuramic acid (product #19023, Sigma Aldrich), n-acetyl-D-glucosamine (product #A4106, Sigma Aldrich), N-Acetyl-2,3-dehydro-2-deoxyneuraminic acid (product #D9050, Sigma Aldrich), 3-hydroxy-DL-kynurenine (product #27778-25, Cayman Chemical), N1,N12-Diacetylspermine (product #17918, Cayman Chemical). A feature table was generated by averaging technical replicates and merging all datasets to process the multiple batches. Targeted metabolite abundances were normalized using a metabolite-specific adjustment method, where each abundance value was divided by the median of the QCs in the respective batch (77).

### Untargeted Metabolomic Analysis

Files were converted to compatible extensions using MSConvert (78). Complementary untargeted analysis was performed and XCMS (76) was used for feature detection, retention time correction, and alignment across samples. Data pre-processing involved selecting features present in at least 80% of samples, removing those with QC Relative Standard Deviation (QC-RSD) > 30%, averaging technical replicates, merging feature datasets using MetabCombiner (79), and applying softImpute (80) in R (81), version 4.5.0. Posterior probabilities were estimated by correcting for batch effect with ComBat (82) then applying RRmix (83) in R to identify differentially abundant metabolites while adjusting for additional unwanted variation.

### Molecule Identification

Collision-induced dissociation (CID) tandem mass spectrometry (MS/MS) was employed to elucidate the structural identities of metabolites identified by the untargeted analysis.

MS/MS fragmentation analysis used two complementary tools to support metabolite identification. The CFM-ID (23) was used to generate predicted fragmentation spectra based on SMILES-formatted molecular structures. These in silico spectra were then compared to the experimental MS/MS spectra to assess the plausibility of candidate metabolite annotations.

Additionally, SIRIUS (22) (version 5) was employed to perform MS/MS-based feature annotation by reconstructing fragmentation trees and scoring candidate molecular formulas and structures. SIRIUS uses fragmentation pattern analysis and isotope pattern matching to determine the most probable chemical structure for a given MS/MS feature.

Fragmentation spectra of pure chemical standards were also acquired and used to complement in silico predictions, aiding in establishing the most probable chemical identifications. Pure chemical standards were purchased: kynurenic acid (product #K3375-250mg, Sigma-Aldrich), and 5-hydroxy-L-tryptophan (product #H9772-100mg, Sigma-Aldrich).

### Cytokine assays

The primary cytokine of interest was IL-6. IL-6 was quantified as part of a multiplex assay employed to optimize data collection from limited-volume serum samples. Following the manufacturer’s instructions the ProcartaPlex™ Human Th1/Th2 Cytokine Panel kit (Thermo Fisher) was used to obtain serum concentrations of GM-CSF, IFN gamma, IL-1 beta, IL-2, IL-4, IL-5, IL-6, IL-12p70, IL-13, IL-18, TNF alpha. Data were acquired using the MAGPIX NxTAG® Luminex system (Diasorin) with xPONENT® Software Solutions for Luminex® Systems. Initial data processing was performed using the Invitrogen ProcartaPlex Analysis App via Thermo Fisher’s Connect platform, and further analysis was conducted in R (80) following normalization steps published elsewhere (84).

### Statistical Analysis

Baseline demographic variables in PTB cases and non-TB controls were presented with counts (percentages) for categorical variables and medians (interquartile ranges, IQR) for numeric variables. Metabolite abundances, as well as cytokine levels, were log_2_-transformed before downstream analyses to normalize distributions. Their levels were compared between groups using the Mann-Whitney test (or the Kruskal-Wallis test for comparisons involving more than two groups).

Multiple hypotheses were not adjusted for the targeted metabolite analysis, as the metabolites were pre-specified based on findings from a prior non-PWH study. A significance threshold of *P <* 0.05 was applied. RRmix(83) - a Bayesian mixture model-was used for the untargeted metabolomics differentially abundant analysis with a significance threshold based on posterior probability > 60%.

Spearman correlation analysis assessed associations between metabolite abundances and cytokine levels (CRP and IL-6), accounting for potential non-linear relationships. To correct for multiple comparisons across the correlation tests, the Holm-Bonferroni method was applied.

All analyses were conducted in the R programming environment (81). Receiver operating characteristic (ROC) curve and area under the curve (AUC) analyses were performed using the pROC package (85), which also implements DeLong’s test (26)—a non-parametric method for comparing the AUCs of two correlated ROC curves. Data wrangling was conducted using dplyr (86), and visualizations were created using ggplot2 (87). Linear mixed-effects models were fitted using lme4 (88), with estimated marginal means computed via the emmeans package (89).

## Data Availability

Supporting data are available with this publication and can be found in the Data Values file and the Supplementary Material. The supporting data include generated targeted and untargeted metabolomic datasets, cytokine values, a covariate table, and underlying values for figures.

## Ethical Considerations

The biospecimen and data handling in this research were obtained from a previous study that strictly adhered to privacy and confidentiality standards, as approved by the institutional review boards (IRB) of the GHESKIO center in Port au Prince, Haiti, Weill Cornell Medical College, and Mass General Brigham. All participants provided written informed consent prior to their inclusion in the clinical cohort study and consented to the use of specimens for future research related to TB.

## Author Contributions

AD, DF, and KR conceptualized the study and designed the methodology. AD performed the experiments and collected the LC-MS and cytokine data. SK, ND, PS, VR, and JP contributed to the collection and provision of clinical samples and clinical data. KR provided the necessary reagents and equipment. Data analysis was carried out by AD, ML, and SB, and AD, ML, SB, DF, SK, ND, KD, and KR performed data interpretation. AD drafted the manuscript, with critical revisions from KD, ML, DF, SK, and KR. All authors reviewed and approved the final version of the manuscript.

## Acknowledgements

This project was supported by the Tri-Institutional Tuberculosis (TB) Research Advancement Center (TRAC) at Weill Cornell Medicine, Memorial Sloan Kettering Cancer Center, and Rockefeller University through the Developmental Project Award (Grant P30AI168433 to Dr. Andrea Doltrario) and by the NIH Tuberculosis Research Units (TBRU) Network grant U19AI162584. Access to human samples would not have been possible without the partnership with the Weill Cornell Center for Global Health, in collaboration with the dedicated staff and participants of the SDART-TB clinical trial, along with the entire team at GHESKIO, Haiti. We acknowledge Maureen Ward for support with urine sample processing and cytokine panel testing. Finally, we thank the TB research community at Weill Cornell Medicine and Tri-I institutes for their valuable insights and support. Partial results from this study were presented at the World Conference on Lung Health 2024 of the International Union Against Tuberculosis and Lung Disease (The Union), Bali, Indonesia, 12 – 16 November 2024.

