## Supplementary Material for "Urine metabolomic biomarkers linked to C-reactive protein-interleukin-6 axis in persons living with HIV and tuberculosis"

Table S1: Multivariate regression model results for urinary metabolites in the full cohort (*n* =298)[A].

| Metabolites | Predictor | USC_B | SE | *P* | CI_Lower | CI_Upper |
|---|---|---|---|---|---|---|
| **m/z 115.0498** | **mTB** | 0.344564414 | 0.149835006 | 0.0222106 | 0.049609114 | 0.639519715 |
| | bmi | -0.015080495 | 0.016297332 | 0.3555941 | -0.047162345 | 0.017001356 |
| | cd4 | -0.000172235 | 0.000260908 | 0.5097133 | -0.000685841 | 0.000341371 |
| **UPA** | **mTB** | 0.39882496 | 0.168775871 | 0.0188140 | 0.066583923 | 0.731065997 |
| | bmi | -0.01591465 | 0.018357501 | 0.3867286 | -0.052052015 | 0.020222715 |
| | cd4 | -0.000172453 | 0.00029389 | 0.5578174 | -0.000750985 | 0.000406079 |
| **N-Acetylhexosamine** | **mTB** | 0.078466617 | 0.081303192 | 0.3353279 | -0.081581478 | 0.238514712 |
| | bmi | -0.00500403 | 0.008843228 | 0.5719453 | -0.022412224 | 0.012404164 |
| | cd4 | 1.55E-05 | 0.000141573 | 0.9128645 | -0.000263186 | 0.000294198 |
| **3-HK** | **mTB** | 0.41702982 | 0.281121367 | 0.1390874 | -0.136367141 | 0.970426781 |
| | bmi | -0.015189148 | 0.030577154 | 0.6197592 | -0.075381315 | 0.045003019 |
| | cd4 | 8.95E-05 | 0.000489517 | 0.8550374 | -0.000874115 | 0.001053146 |
| **Unknown 2** | **mTB** | -0.159791888 | 0.1678283 | 0.3418656 | -0.490167598 | 0.170583822 |
| | bmi | 0.000883336 | 0.018254435 | 0.9614400 | -0.035051141 | 0.036817812 |
| | cd4 | 0.000235495 | 0.00029224 | 0.4210309 | -0.000339789 | 0.000810778 |
| **Neopterin** | **mTB** | -0.019637533 | 0.176841474 | 0.9116600 | -0.367755982 | 0.328480917 |
| | bmi | -0.034171561 | 0.019234785 | 0.0767358 | -0.072035888 | 0.003692767 |
| | cd4 | -0.000130827 | 0.000307934 | 0.6712730 | -0.000737006 | 0.000475352 |
| **Sialic Acid 1** | **mTB** | 0.038947937 | 0.090143193 | 0.6660273 | -0.138502002 | 0.216397875 |
| | bmi | -0.00677018 | 0.009804741 | 0.4904555 | -0.026071147 | 0.012530787 |
| | cd4 | -0.000121581 | 0.000156966 | 0.4392552 | -0.000430574 | 0.000187413 |
| **Sialic Acid 2** | **mTB** | 0.054315124 | 0.08496194 | 0.5231622 | -0.112935343 | 0.22156559 |
| | bmi | -0.010382466 | 0.009241184 | 0.2621947 | -0.028574052 | 0.007809119 |
| | cd4 | 2.07E-05 | 0.000147944 | 0.8889345 | -0.000270554 | 0.000311913 |
| **Sialic Acid 3** | **mTB** | -0.217178491 | 0.176897961 | 0.2205963 | -0.565408138 | 0.131051155 |
| | bmi | -0.044887573 | 0.019240929 | *0.0203658* | -0.082763996 | -0.007011151 |
| | cd4 | -6.92E-05 | 0.000308033 | 0.8224656 | -0.000675553 | 0.000537193 |
| **DiAcSpm** | **mTB** | 0.138222804 | 0.186438391 | 0.4590853 | -0.228787502 | 0.50523311 |
| | bmi | 0.00689994 | 0.020278627 | 0.7339201 | -0.033019227 | 0.046819107 |
| | cd4 | 3.32E-06 | 0.000324645 | 0.9918518 | -0.000635757 | 0.000642394 |
| **m/z 115.0498** | **cTB+mTB** | 0.356024937 | 0.13598744 | 0.0093008 | 0.088392722 | 0.623657152 |
| | bmi | -0.013402047 | 0.015918971 | 0.4005330 | -0.044731627 | 0.017927533 |
| | cd4 | -0.000228865 | 0.000254583 | 0.3693985 | -0.0007299 | 0.00027217 |
| **UPA** | **cTB+mTB** | 0.41324321 | 0.152821816 | 0.0072483 | 0.112479837 | 0.714006583 |
| | bmi | -0.013682256 | 0.017889637 | 0.4449954 | -0.048890238 | 0.021525725 |
| | cd4 | -0.000242811 | 0.000286098 | 0.3967409 | -0.000805871 | 0.00032025 |
| **N-Acetylhexosamine** | **cTB+mTB** | 0.105109064 | 0.07329252 | 0.1526054 | -0.03913543 | 0.249353558 |
| | bmi | -0.00405491 | 0.008579774 | 0.6368401 | -0.020940469 | 0.012830648 |
| | cd4 | -2.66E-07 | 0.000137211 | 0.9984567 | -0.000270306 | 0.000269775 |
| **3-HK** | **cTB+mTB** | 0.586024818 | 0.258746147 | 0.0242489 | 0.076795403 | 1.095254232 |
| | bmi | -0.011086625 | 0.030289359 | 0.7146106 | -0.070698072 | 0.048524822 |
| | cd4 | -8.34E-06 | 0.0004844 | 0.9862783 | -0.000961669 | 0.000944993 |
| **Unknown 2** | **cTB+mTB** | -0.171989626 | 0.152888407 | 0.2615338 | -0.472884055 | 0.128904802 |
| | bmi | 0.006489721 | 0.017897433 | 0.7171596 | -0.028733603 | 0.041713044 |
| | cd4 | 0.00016678 | 0.000286223 | 0.5605466 | -0.000396526 | 0.000730086 |
| **Neopterin** | **cTB+mTB** | 0.097227666 | 0.161247537 | 0.5469925 | -0.220118079 | 0.414573411 |
| | bmi | -0.029701259 | 0.01887597 | 0.1166795 | -0.066850408 | 0.007447889 |

| | | | | | | |
|---|---|---|---|---|---|---|
| | cd4 | -0.000192149 | 0.000301872 | 0.5249308 | -0.000786253 | 0.000401956 |
| **Sialic Acid 1** | **cTB+mTB** | 0.080705525 | 0.081626456 | 0.3236143 | -0.079940702 | 0.241351753 |
| | bmi | -0.005236927 | 0.009555362 | 0.5840655 | -0.024042506 | 0.013568653 |
| | cd4 | -0.00013597 | 0.000152813 | 0.3743111 | -0.000436717 | 0.000164776 |
| **Sialic Acid 2** | **cTB+mTB** | 0.091884032 | 0.076990457 | 0.2336565 | -0.059638245 | 0.243406308 |
| | bmi | -0.009369922 | 0.009012662 | 0.2993615 | -0.027107433 | 0.008367589 |
| | cd4 | 4.37E-06 | 0.000144134 | 0.9758314 | -0.000279295 | 0.000288036 |
| **Sialic Acid 3** | **cTB+mTB** | -0.084628747 | 0.16086483 | 0.5992250 | -0.4012213 | 0.231963807 |
| | bmi | -0.04271837 | 0.018831169 | 0.0240242 | -0.079779348 | -0.005657391 |
| | cd4 | -4.68E-05 | 0.000301156 | 0.8765021 | -0.000639536 | 0.000545852 |
| **DiAcSpm** | **cTB+mTB** | 0.171061026 | 0.168826576 | 0.3117810 | -0.161200766 | 0.503322818 |
| | bmi | 0.008916195 | 0.019763188 | 0.6522128 | -0.029979057 | 0.047811446 |
| | cd4 | -4.80E-05 | 0.000316061 | 0.8794975 | -0.000669987 | 0.00057407 |

[A]Three participants without available CD4+ T-cell counts were excluded from this analysis. This table presents results from multivariate linear regression models evaluating associations between urinary metabolite levels and participant groups, adjusted for body mass index (BMI) and CD4+ T-cell count—covariates that significantly differed between groups. Two comparisons were assessed: (1) mTB participants versus non-TB controls, and (2) combined mTB and cTB participants versus non-TB controls. P-values (P) $< 0.05$ were considered statistically significant. USC B: unstandardized regression coefficient; SE: standard error of the regression coefficient; CI: confidence interval; UPA: ureidopropionic acid; 3-HK: 3-hydroxykynurenine; DiAcSpm: N1,N12-Diacetylspermine; mTB: microbiological TB group; cTB: clinical TB group. BMI: body mass index.

**Figure S1.**

**Figure S1. Chromatography associated with untargeted metabolomic analysis. A)** Chromatographic representation of peaks corresponding to the fragments and $^{13}C$ isotope of the parent ion m/z 221.0925, predicted to be a hydroxytryptophan isoform. Numbers above the peaks represent the m/z of each detected peak, followed by its relative height (%) to the parent ion m/z 221.0925. **B)** Chromatographic comparison of a QC sample spiked with a pure 5-HTP chemical standard (purple line) versus a non-spiked QC sample (gray line). As shown in the figure, the urinary HTP (u-HTP) isoform—identified as a significant feature in the untargeted analysis—does not align with the retention time of the 5-HTP chemical standard, confirming that the detected metabolite is distinct. Retention times in both (A) and (B) exhibit slight variations compared to those reported in Table 3 of the main manuscript. These differences are expected due to inherent variability in chromatography conditions across different runs. **(C)** The table presents peak areas, heights, and experimentally observed relative abundances. The observed relative abundances for $^{13}C$ isotope peaks, determined by area or height, closely align with the expected calculated values (12.9% and 11.4% for HTP and KYNa, respectively). The data presented here were obtained from a quality control (QC) pooled urine sample. [A,B] Features predicted to be $^{13}C$ isotopes from their respective parent ions.

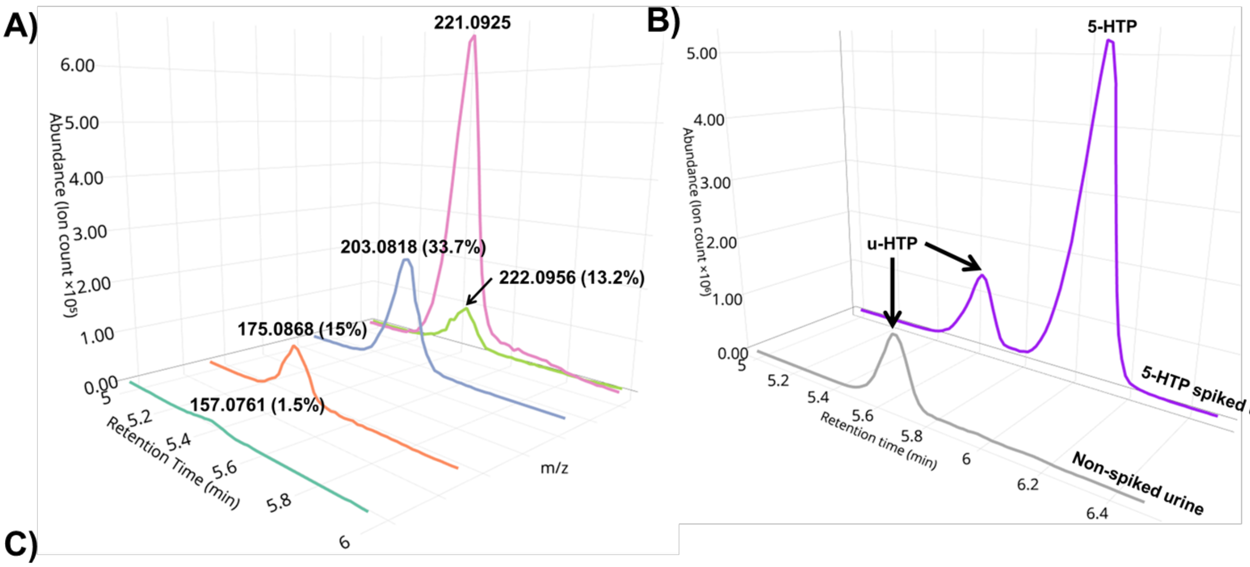


C)

| m/z | Peak area | Peak height | Relative area | Relative height |
|---|---|---|---|---|
| **HTP - C11H12N2O3** | | | | |
| **221.0925** | 5,623,260.49 | 674,024.75 | 100.0% | 100.0% |
| **222.0956**[A] | 663,628.56 | 89,259.54 | 11.8% | 13.2% |
| **203.0818** | 1,860,017.46 | 227,359.67 | 33.1% | 33.7% |
| **175.0828** | 814,827.57 | 101,316.18 | 14.5% | 15.0% |
| **157.0761** | 79,387.24 | 10,273.87 | 1.4% | 1.5% |
| **KYNa - C10H7NO3** | | | | |
| **190.0503** | 3,428,739.88 | 1,250,477.33 | 100.0% | 100.0% |
| **191.0535**[B] | 428,093.44 | 157,289.51 | 12.5% | 12.6% |

**Table S2. Characteristics of participants included in the serum analysis stratified by groups**

| | Non-TB (*n* =32) | cTB (*n* =16) | mTB (*n* =32) | *P* [A] |
|---|---|---|---|---|
| **Age (yr), median [IQR]** | 38 [33, 41.8] | 40.5 [31.5, 45] | 38 [34.2, 43] | 0.999 |
| **BMI ($kg/m^2$), median [IQR]** | 21.3 [20, 22.8] | 19.7 [17.1, 21.1] | 19.2 [18, 20.7] | <0.001 |
| **Sex, n(%)** | | | | 1 |
| Female | 10 (31.2%) | 5 (31.2%) | 10 (31.2%) | |
| Male | 22 (68.8%) | 11 (68.8%) | 22 (68.8%) | |
| **CD4+ T count(cells/$mm^3$), median [IQR]** | 288 [168.2, 399] | 196.5 [62.5, 309.2] | 167.5 [91.2, 289.8] | 0.067 |
| Missing values | 0 | 0 | 2 (6.2%) | |
| **CRP (mg/dL), median [IQR]** | 2.5 [1.5, 10.2] | 49.9 [10.2, 125.8] | 33.3 [4.9, 88] | 0.0014 |
| **Prevalence of reported symptoms, n(%)** | | | | |
| Cough | 11 (34.4%) | 14 (87.5%) | 23 (71.9%) | <0.001 |
| Fever | 10 (31.2%) | 10 (62.5%) | 23 (71.9%) | 0.004 |
| Night Sweats | 7 (21.9%) | 5 (31.2%) | 13 (40.6%) | 0.274 |
| Weight Loss | 30 (93.8%) | 15 (93.8%) | 31 (96.9%) | 0.823 |
| **Chest X-ray[B], n(%)** | | | | 0 |
| Abnormal | 3 (9.4%) | 15 (93.8%) | 15 (46.9%) | 0 |
| Normal | 29 (90.6%) | 1 (6.2%) | 17 (53.1%) | |

[A] Reported P-values reflect overall comparisons among the three groups using the multigroup statistical test. Numeric variables were tested using ANOVA, and categorical variables were analyzed with the Kruskal-Wallis test for multiple groups. P-values (P) <0.05 were considered statistically significant.[B] Attending physician chest X-ray assessment**.** TB: tuberculosis; PTB: pulmonary tuberculosis; cTB: clinical TB group; mTB: microbiological TB group. BMI: body mass index; CRP: C-reactive protein.

**Table S3. Th1/Th2 cytokine panel group comparison results**

| | Non-TB (*n* =32) | mTB (*n* =32) | cTB+mTB (*n* =48) | ***P*** |
|---|---|---|---|---|
| | | MFI (median) | | |
| **mTB** | | | | |
| **IL6** | 7.00 | 8.25 | NA | 0.022 |
| **GM-CSF** | 5.50 | 5.50 | NA | 0.654 |
| **IFN gamma** | 11.50 | 15.00 | NA | 0.320 |
| **IL-1 beta** | 3.87 | 4.00 | NA | 0.183 |
| **IL-2** | 5.50 | 5.00 | NA | 0.793 |
| **IL-4** | 5.00 | 5.50 | NA | 0.658 |
| **IL-5** | 5.12 | 5.50 | NA | 0.432 |
| **IL-12p70** | 7.00 | 7.12 | NA | 0.957 |
| **IL-13** | 5.50 | 5.50 | NA | 0.972 |
| **IL-18** | 49.50 | 65.78 | NA | 0.240 |
| **TNF alpha** | 7.37 | 7.50 | NA | 0.714 |
| **cTB+mTB** | | | | |
| **IL6** | 7.000 | NA | 8.000 | 0.006 |
| **GM-CSF** | 5.500 | NA | 5.500 | 0.602 |
| **IFN gamma** | 11.500 | NA | 13.491 | 0.536 |
| **IL-1 beta** | 3.873 | NA | 4.000 | 0.311 |
| **IL-2** | 5.500 | NA | 5.000 | 0.804 |
| **IL-4** | 5.000 | NA | 5.500 | 0.777 |
| **IL-5** | 5.123 | NA | 5.500 | 0.625 |
| **IL-12p70** | 7.000 | NA | 7.000 | 0.640 |
| **IL-13** | 5.500 | NA | 5.500 | 0.947 |
| **IL-18** | 49.499 | NA | 53.722 | 0.596 |
| **TNF alpha** | 7.374 | NA | 7.500 | 0.878 |

Group comparisons were performed using Mann-Whitney U test, with statistical significance set at $P < 0.05$. PTB: pulmonary tuberculosis; mTB: microbiological TB group; cTB: clinical TB group; MFI: median fluorescence intensity; *P* : p-value.
